# A germline heterozygous *POLQ* nonsense mutation causes hereditary colorectal cancer

**DOI:** 10.1101/2024.01.13.23299913

**Authors:** Ning Xu, Deng-Feng Zhang, Kexin Yang, Yu Fan, Fengchang Huang, Junyu Ren, Rui Bi, Yu Li, Maosen Ye, Min Xu, Yongchun Zhou, Wenhui Li, Xiaoxiao Shi, Yubo Wei, Chao Zhang, Yong-Gang Yao, Wen-Liang Li

## Abstract

The causal genes for a large proportion of hereditary colorectal adenomas and early-onset colorectal cancer (CRC) remain to be identified. Here, we identified a germline heterozygous stop-gain mutation p.Arg1953X (rs150312701) of the *POLQ* (DNA Polymerase Theta) gene, which is co-segregated with disease status, by whole-exome sequencing of twelve hereditary CRC pedigrees. The mutation was validated in an independent pedigree, resulting in ten p.Arg1953X carriers from two CRC families. Mechanically, the heterozygous nonsense mutation led to compensated overexpression of the mRNA with wild-type *POLQ* allele under DNA damage stress, resulting in hyperactivation of the error-prone theta mediated end-joining (TMEJ) DNA repair pathway, which enables the survival of mutation-enriched cells. Concordantly, tumor tissues from p.Arg1953X mutation carriers showed microsatellite instability and hypermutation, and were resistant to radiotherapy. We found that an FDA-approved antibiotic Novobiocin inhibits the *POLQ*-mediated TMEJ pathway, eliminates the p.Arg1953X mutation-related resistance to DNA damage, finally benefits tumor radiotherapy. Collectively, we defined a POLQ-mutated CRC type and suggested for a mutation-based potential target therapeutic strategy.

---

Mendelian cancer syndromes account for approximately 5% of all colorectal cancer (CRC) cases^1^. The most recognized of these syndromes is Lynch syndrome (LS), characterized by early-onset CRC or endometrial cancer^2^. Familial adenomatous polyposis (FAP) and *MUTYH*-associated polyposis (MAP), primarily predispose individuals to multiple adenomas, the benign precursors to many CRCs^3,4^. Other less common Mendelian syndromes associated with CRC include juvenile polyposis syndrome (JPS), *PTEN* hamartoma tumor syndrome (PHTS), Peutz–Jeghers syndrome (PJS), polymerase proofreading-associated polyposis (PPAP), and *NTHL1*-associated polyposis (NAP)^5–7^. Pathogenic genes for these known syndromes serve different functions, with four DNA mismatch repair genes (*MSH2*, *MLH1*, *MSH6*, and *PMS2*), two base excision repair genes (*MUTYH* and *NTHL1*), and two DNA proofreading repair genes (*POLD1* and *POLE*). These genes contribute to tumorigenesis by impacting DNA repair, which are fundamental for maintaining genome stability and whose defects are linked to cancer predisposition^8^. However, the etiology of many familial CRC cases remains unexplained.

We herein analyzed a dozens of families likely to harbor novel high-penetrance germline CRC driven genes, as known causal mutations were absent in these families. Employing a two-stage strategy encompassing whole-exome sequencing (WES) and Sanger sequencing of both familial and sporadic CRC cases, we discerned a co-segregation of a nonsense POLQ mutation p.Arg1953X with disease phenotype. Utilizing *in vitro* and *in vivo* approaches, we uncovered the molecular mechanism by which the mutation leads to tumorigenesis. We further provided a candidate drug, Novobiocin, for targeting POLQ-associated cancers.

## Results

### Identifying *POLQ* as a new pathogenic gene for hereditary colorectal cancer

In the discovery phase of our study, we performed whole exome sequencing (WES) of 43 subjects from 12 pedigrees (**Fig. 1a-b**, **Supplementary Fig. S1**), of which the probands were clinically diagnosed with hereditary colorectal cancer (CRC) and at least one first-degree relative suffering from polyposis or CRC. For each family, at least two affected individuals and one healthy control were sequenced. Initial screening was carried out for the presence of deleterious variants in established pathogenic genes, and we identified an *MSH2* variant (encoding a p.Arg645* alteration) in family LS6 (**Supplementary Table 1**).

**Figure 1.**
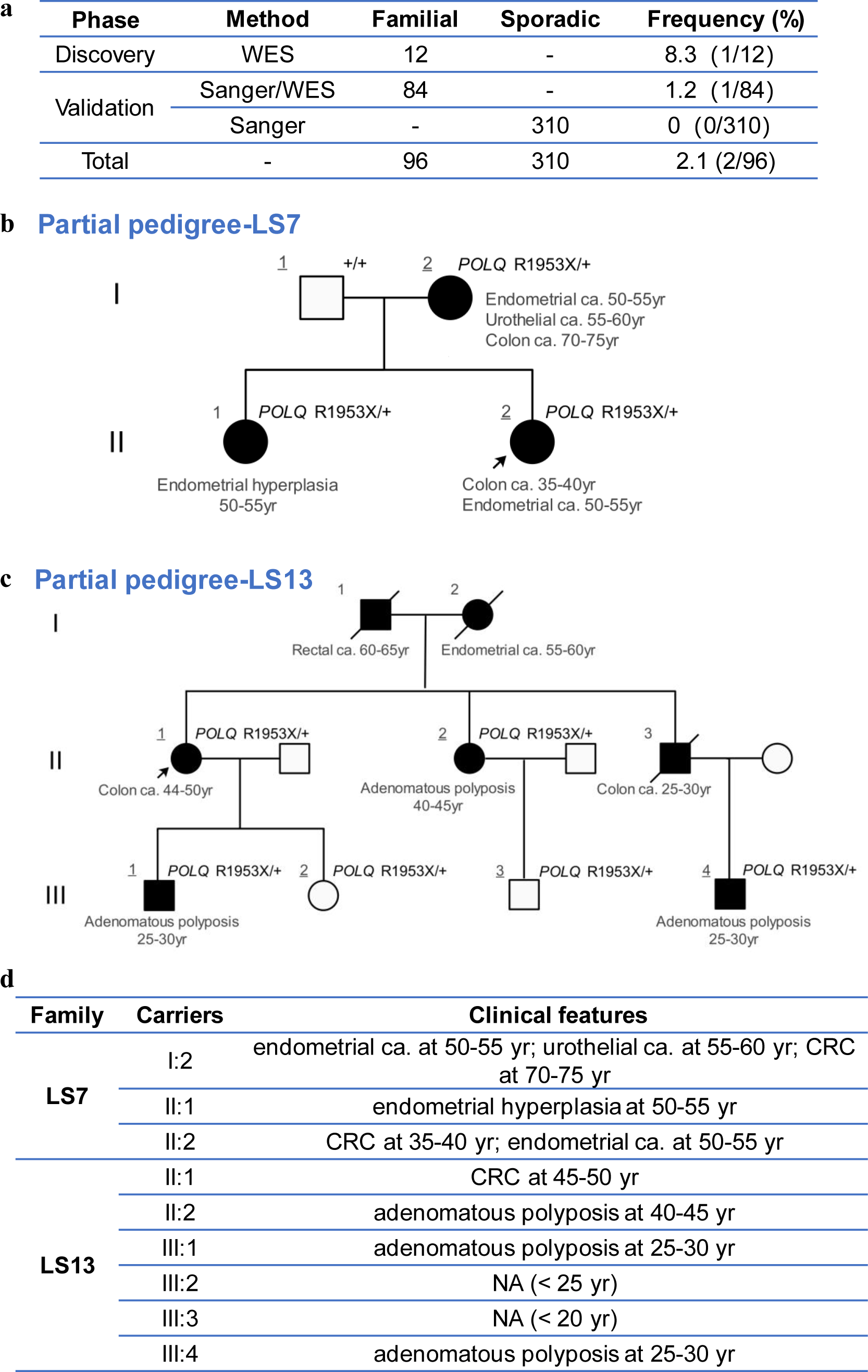
Identification of pathogenic germline POLQ mutation in pedigrees with colorectal cancer. **a**, Whole exome sequencing was conducted in 12 pedigrees with colorectal cancer (CRC). The *POLQ* mutation was then validated in 84 familial CRC cases and 310 sporadic CRC cases. **b**, three patients are *POLQ* Arg1953X carriers in pedigree LS7 in the discovery stage. Individuals with underlined number were whole exome sequenced; ca., cancer, with the following numbers refer to the age at first presentation. Partial pedigree was shown to exclude identifying information. **c**, the POLQ Arg1953X mutation was detected in one proband of the validation cohort, and the mutation was further screened in its family (pedigree LS13). Partial pedigree was shown to exclude identifying information. **d**, Clinical features of the mutation carriers. CRC, colorectal cancer; yr, years-old; NA, not available; Numbers represent age (in years) of onset of clinical feature present at the time of diagnosis.

To uncover potentially novel pathogenic mutations and causal genes, we conducted comprehensive genomic annotation and co-segregation analyses in remaining pedigrees with no known pathogenic mutations. In those rare damaging variants showing co-segregation with disease status in the LS7 pedigree (**Supplementary Table 2**), a germline mutation c.5857C>T (rs150312701; NM_199420 p.Arg1953X, which is a premature termination codon (PTC) mutation) in the *POLQ* (DNA Polymerase Theta) gene showed the most deleterious effect (CADD score = 42). Further Sanger sequencing validated the presence and co-segregation of the mutation in the LS7 pedigree (**Fig.1b**). Note that this mutation was reported in previous studies on colorectal cancer (CRC) organoid cultures and astrocytic tumors, but was documented as a heterozygous somatic mutation in these reports^9,10^.

In order to validate the association of POLQ p.Arg1953X with CRC and to clarify the mutation frequency, we genotyped this mutation in 84 Chinese individuals diagnosed as the proband of hereditary CRC pedigrees. All these individuals came from families with an overrepresentation of familial history of CRC, adenomas, and other early-onset cancer across five medical institutions (**Supplementary Table 3**). 310 individuals with sporadic colorectal cancer were also screened for p.Arg1953X. We identified a genetically unrelated case (LS13, II:2) to family LS7 harboring the p.Arg1953X mutation, whereas no mutation was identified in sporadic CRC cases (**Fig.1a, c-d**). Genotyping of additional members of the LS13 family revealed that most carriers of the p.Arg1953X mutation developed colorectal tumors, affirming the co-segregation of the p.Arg1953X with CRC status. With 2 out of 96 scrutinized families, this mutation was observed at a reasonably high frequency of 2.1% in Chinese patients with a family history of CRC (**Fig 1a**).

The phenotypes discerned in both pedigrees carrying p.Arg1953X exhibited a dominantly inherited mode, aligning with a high-penetrance predisposition towards colorectal adenoma and carcinoma, with a propensity to predispose extra-colonic tumors. Among the ten p.Arg1953X carriers of the two families, three individuals developed extra-colonic tumors, and two had developed multiple primary cancers in the LS7 family. The cases of the LS13 family predominantly exhibited colorectal tumors (**Fig. 1d**).

### Isogenic cell lines with POLQ p.Arg1953X mutation have higher viability in response to DNA-damage

To investigate whether the the POLQ p.Arg1953X promotes the pathogenesis of tumor, we introduced this mutation into 293T and HCT116 cell lines (*POLQ*^+/R1953X^ and *POLQ*^R1953X/R1953X^), respectively, using the CRISPR/Cas9-mediated homologous recombination (**Supplementary Fig. S1**). To check whether the mutation leads to a loss-of function of the protein, we also established a HCT116 cell line with POLG knockout (*POLQ*^-/-^), as a control for comparison with p.Arg1953X cells (**Supplementary Fig. S1**).

The *POLQ* gene has been found to be specifically involved in the repair of DNA double-strand breaks (DSBs)^11,12^, we thus examined the viability of *POLQ* mutant cells in response to DNA-damage stress. We found that *POLQ*^+/R1953X^ cells were significantly less sensitive to Ionizing Radiation (IR) or ultraviolet (UV) induced DNA damage compared with *POLQ*^+/+^ cells (**Fig. 2a**). Consistently, the survival assay showed that increasing concentrations of hydroxyurea (HU) (0-0.5mM), ETOPOSIDE (0-5uM), UV (0-15J/m^2^) or IR (0-3Gry) inhibited the number of clones of these cells in a dose-dependent manner, but their effect on *POLQ*^+/R1953X^ cells were significantly lower than those on *POLQ*^+/+^ cells (**Fig. 2b-d**). Furthermore, 293T cells with the p.Arg1953X mutation had a promoted cell cycle progression after DNA-damage, as indicated by increased DNA-synthesis (S) phase which is most vulnerable period of the cell cycle during genotoxic stresses (**Fig. 2e**). In line with this observation, the mutation significantly promoted DNA synthesis after DNA-damage, as measured by 5-ethynyl-2’-deoxyuridine (EdU) incorporation assay (**Fig. 2f-g**). Consequently, the cells with the p.Arg1953X mutation are resistant to apoptosis (**Fig. 2h-i**).

**Figure 2.**
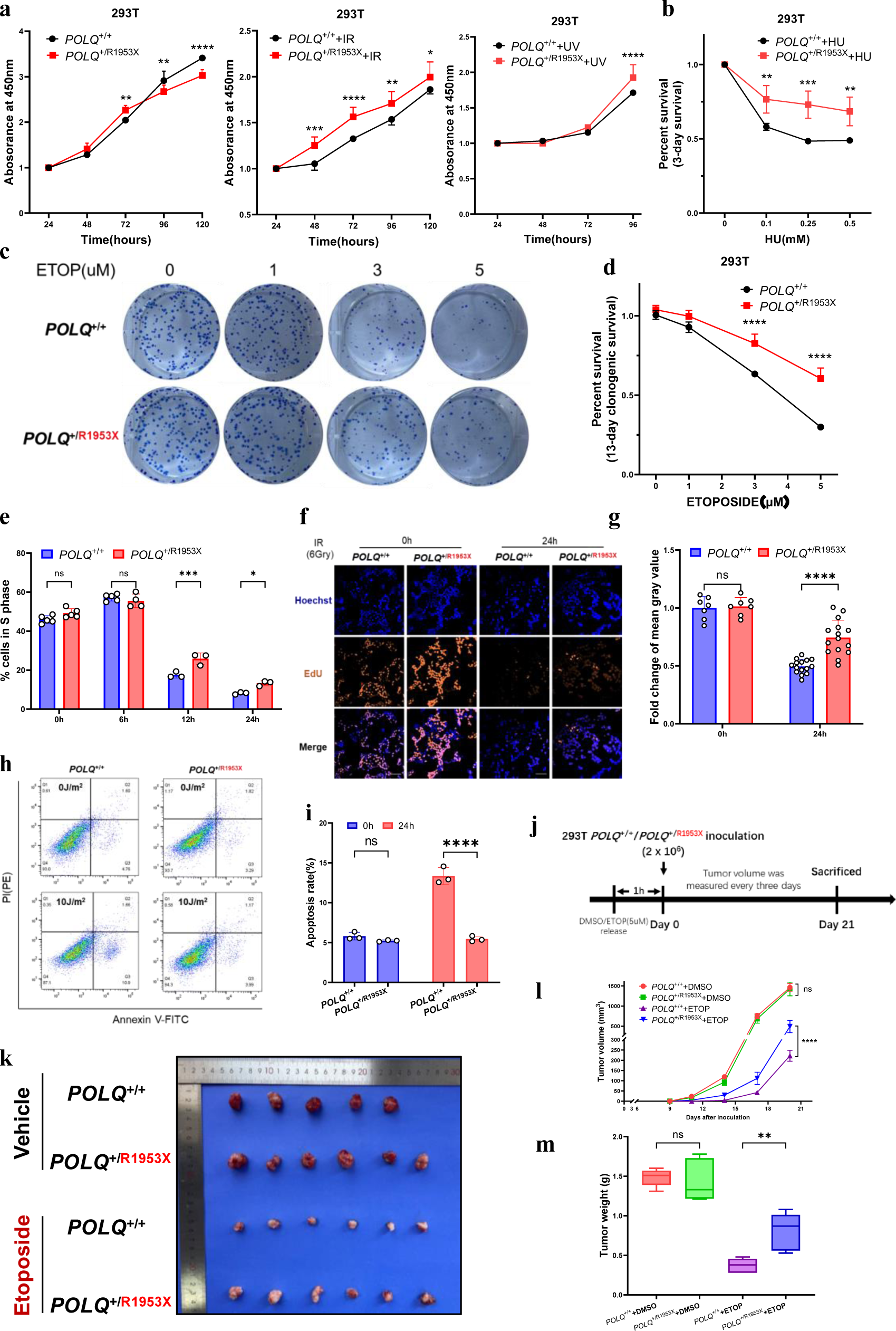
Cells with *POLQ* Arg1953X mutation escape from apoptosis and survive better under DNA-damage stress. **a**, CCK8 assay showed that 293T cells with heterozygous *POLQ* Arg1953X mutation (*POLQ*^+/R1953X^) have higher viability when exposed to UV (10 J/m^2^) or Ionizing radiation (IR, 4 Gry) treatments, which induce DNA lesions. **b**, *POLQ*^+/R1953X^ cells survive better than *POLQ*^+/+^ cells under hydroxycarbamide (HU, 0-0.5mM) treatment. **c-d**, *POLQ*^+/R1953X^ cells survive better than *POLQ*^+/+^ cells under ETOPOSIDE (ETOP, 0-5uM) treatment. **e**, there are more *POLQ*^+/R1953X^ 293T cells in the S phase of cell cycle, under the cellular stress and DNA damage induced by IR (6 Gy). Cell cycle distribution was analyzed by flow cytometry and FlowJo software. *P*-value was determined by two-way analysis of variance. **f-g**, Fraction of cycling 293T cells (*POLQ*^+/R1953X^ and *POLQ*^+/+^) measured by EdU incorporation at 0h and 24h after IR (6 Gy) release. The images were taken at ×100 magnification. *P*-value was determined by two-way analysis of variance. **h-i**, *POLQ*^+/R1953X^ 293T cells resistant to UV-induced apoptosis. Apoptosis was detected by annexin/PI staining and flow cytometry analysis in 293T cells. The cells were cultured for 24h after exposure to UV(10J/m^2^). *P*-value was determined by two-way analysis of variance. **j**, Diagram shows the experimental protocol for DNA-lesion-induced 293T cell xenografting in BALB/c nude mice. **k-m**, Heterozygous *POLQ* R1953X mutation promotes the growth of DNA-lesion-induced 293T cells in vivo. *POLQ*^+/R1953X^ and *POLQ*^+/+^ 293T cells were treated with ETOPOSIDE or DMSO and then xenografted in BALB/c nude mice. Tumor specimens were collected on day 21 after 293T inoculation. Tumor volumes (**l**) and tumor weights (**m**) were measured in different groups (n=6/group). *P*-value was determined by Student’s *t*-test. Data are shown as mean ± SD. \**P* < 0.05; \*\**P* < 0.01; \*\*\**P* <0.001; \*\*\*\**P* < 0.0001 and ns, not significant.

To further confirm the enhanced resistance of *POLQ* p.Arg1953X mutation under replicative stress and DNA-damage *in vivo*, nude mice were inoculated with *POLQ*^+/+^ and *POLQ*^+/R1953X^ 293T cells and HCT116 cells, respectively, which were induced by DNA-damage agent (**Fig. 2j**). Xenograft volume and mouse weight were monitored during the whole course (**Fig. 2j-k**). Heterozygous mutation promoted xenograft growth and increased tumor weight (**Fig. 2k-m**). These results suggest that the POLQ p.Arg1953X mutation is associated with abnormally enhanced resistance to DNA-damage, which finally promotes tumorigenesis.

### POLQ p.Arg1953X mutation leads to overactivation of *POLQ* level under stress

The prevailing hypothesis is that PTC mutation might induce protein haploinsufficiency or a truncate peptide. We observed that the total *POLQ* mRNA expression level in mutated individuals was reduced in blood compared with that of individuals without the mutation (**Supplementary Fig. S2a**). Similarly, we observed a significantly reduced expression of *POLQ* mRNA in *POLQ*^+/R1953X^ (293T and HCT116) cell lines when compared with *POLQ*^+/+^ cell lines (**Supplementary Fig. S2b**). At the protein level, we observed comparable abundant of POLQ among isogenic cell lines by Western blot analysis using an antibody detecting the N-terminus of POLQ in all comparisons, whereas POLQ protein expression was undetectable in *POLQ*^R1953X/R1953X^ cell lines (**Supplementary Fig. S2c**). We did not detect a truncated POLQ peptide by Western blot analysis using an N-terminus antibody, when compared with wild-type Myc-POLQ and truncated mutant Myc-POLQ^R1953X^ (**Supplementary Fig. S2c**).

To further evaluated whether the single wild-type allele is sufficient to achieve the typical phenotype, we treated isogenic and control cell lines (*POLQ*^+/R1953X^ and *POLQ*^+/+^) with DNA-damage agents and then measured *POLQ* expression levels at different times after treatment. We found that the mRNA expression patterns of *POLQ* in the two genotypes cells were significantly different. Compared with *POLQ*^+/+^ cells, the expression of *POLQ* was increased significantly in *POLQ*^+/R1953X^ cells at 2 hours after DNA-damage exposure to UV or ETOPOSIDE (**Fig. 3a**). RNA-sequencing of HCT116 cell lines with *POLQ*^+/R1953X^, *POLQ*^+/+^ and *POLQ*^-/-^ also showed a significant increase of *POLQ* mRNA level in *POLQ*^+/R1953X^ cellsat 2 hours after IR-induced DNA damage relative to *POLQ*^+/+^ cell lines (**Fig. 3b**).

**Figure 3.**
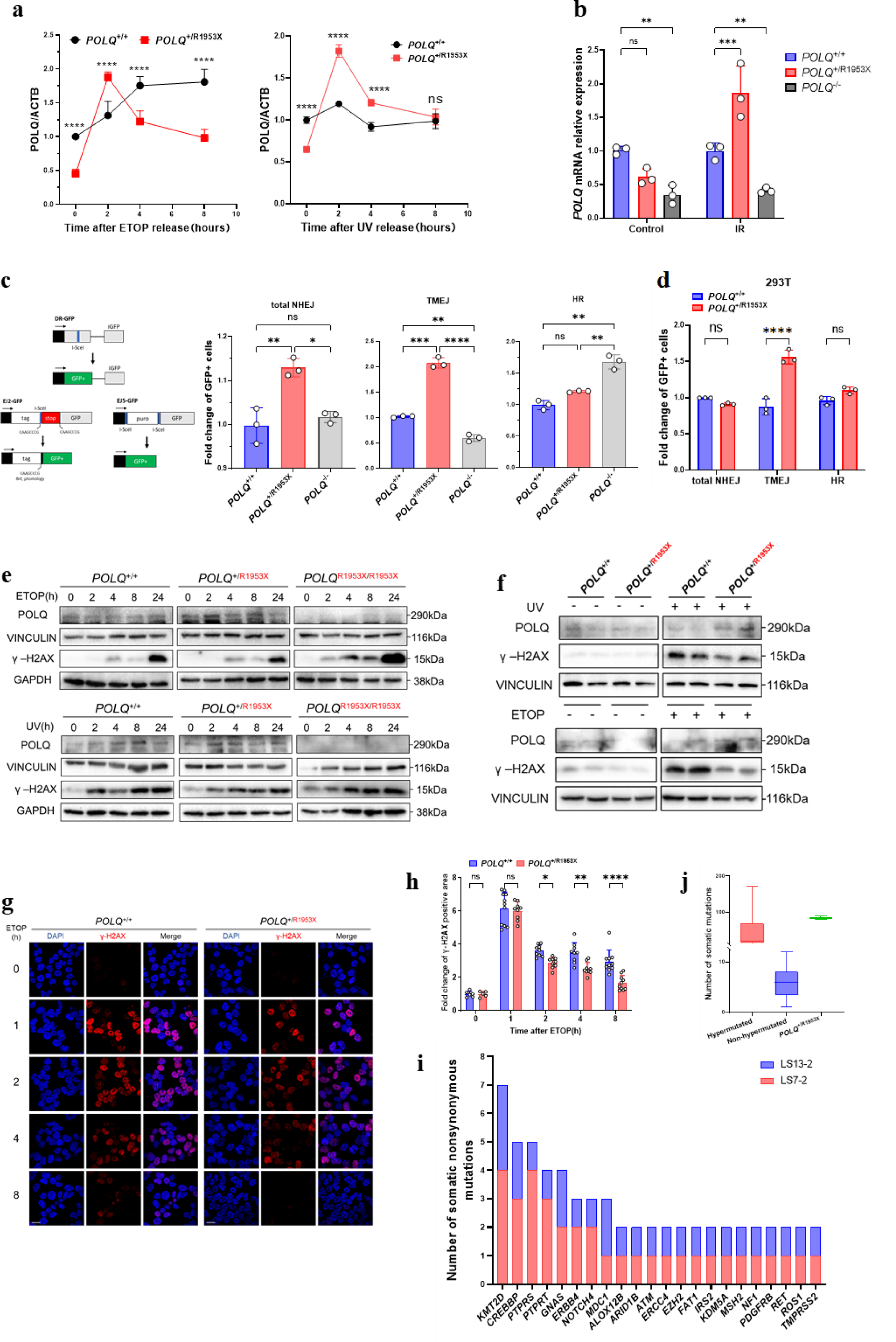
Activated error-prone DNA repair pathway accounts for the tumorigenesis. **a**, *POLQ* mRNA level of *POLQ*^+/R1953X^ cells increased early in response to DNA damage. **b**, *POLQ* expression level increased in *POLQ*^+/R1953X^ HCT116 cell lines 2h after 6 Gy IR induction compared with *POLQ*^+/+^ cells. *P*-value was determined by two-way analysis of variance. **c-d**, Flow cytometric analysis of total-NHEJ, TMEJ and HR efficiency showed that *POLQ* R1953X mutation promoted TMEJ activity in HCT116 (**c**) and 293T (**d**) cells. **e**, Increased *POLQ* protein level and decreased DNA damage in *POLQ*^+/R1953X^ HCT116 cells and 293T cells (f) treated with ultraviolet (UV) or ETOPOSIDE (ETOP), which is time-dependent. VINCILIN, TUBULIN and GADPH were used as the loading control. **g-h**, Reduced ETOP and UV induced DNA damages in *POLQ*^+/R1953X^ 293T cells. DNA double-strand breaks measured by γ-H2AX level. Scale bars, 20µm. Data are shown as mean ± SD from at least three independent biological replicates. *P*-value was determined by two-way ANOVA. \**P* < 0.05; \*\**P* < 0.01; \*\*\**P* <0.001; \*\*\*\**P* < 0.0001 and ns, not significant. **i**, Higher somatic mutation burden in cancer tissues derived from *POLQ* R1953X carriers, with frequently mutated oncogenes shown in (**j**).

### POLQ p.Arg1953X mutation promotes DNA repair via the TMEJ pathway

We then investigated the impact of the overexpression of POLQ by the p.Arg1953X mutation on DNA repair, as the POLQ protein is central in polymerase theta-mediated end joining (TMEJ), a DNA double-strand break (DSB) repair pathway. We measured activity of total nonhomologous end-joining (NHEJ), TMEJ, and homologous recombination (HR) using reporter systems EJ5GFP, EJ2GFP, and DRGFP, respectively (**Fig. 3c**)^13^. The results showed that the p.Arg1953X mutation promoted TMEJ and total NHEJ activity in *POLQ*^+/R1953X^ cells (**Fig. 3c-d**). These results demonstrated that the p.Arg1953X mutation causes overactivation of NHEJ and TMEJ pathway via compensated overexpression of wild-type allele of POLQ, resulting in enhanced DNA repair. Indeed, we found that the ETOP (and UV)-induced DNA damages in *POLQ*^+/R1953X^ 293T and HCT116 cells were decreased compared with *POLQ*^+/+^ cells as indicated by the γ-H2AX level, a marker of DNA damage, as analyzed by both Western blot and Immunofluorescence (**Fig. 3e-h)**. In parallel, the expression of *POLQ* in *POLQ*^+/R1953X^ cells was significantly higher than that of the *POLQ*^+/+^cells with ETOP and UV treatment, as detected by using the N-terminal antibody (**Fig. 3e-f**).

### Sequencing of isogenic cell lines and patient-derived samples reveal a gene signature of tumor with POLQ p.Arg1953X mutation

Since the TMEJ-mediated DNA repair is error-prone, we speculated that such a process might lead to higher mutation burden. We thus sequenced 769 cancer-related genes in the CRCs of two affected individuals from LS7 (II:2) and LS13 (II:2) families and 320 individuals without the POLQ p.Arg1953X mutation. Indeed, we found that the two carriers had high level tumor mutational burden (TMB-H) and microsatellite instability (MSI-H). A total of 89 and 80 nonsynonymous somatic mutations were observed in two affected individuals, respectively (**Fig. 3i, Supplementary Table S4**). Compared with the individuals without the POLQ p.Arg1953X mutation, these numbers are within the range of hypermutated CRCs (**Fig. 3j**), though no mutations were observed in frequently mutated driver genes in hypermutated CRC (**Fig. 3i**) ^14^. And there is a biased mutation pattern toward C:G>T:A transitions and insertions/deletions (In/Del), as usually seen in MMR-deficient cancer cells ^15^. Collectively, these findings showed that cells with POLQ p.Arg1953X mutation might develop with genomic instability, which might drive tumorigenesis and the mutated tumor cells may turn to hypermutation/MSI subtype.

### Introduction of another premature termination codon POLQ p.Q1949X mimic the R1953X-related phenotypes

To verify the PTC-mediated mechanism, we introduced an artificial heterozygous PTC mutation p.Gln1949* in 293T and HCT116 cell lines (*POLQ*^+/Q1949X^ and *POLQ*^Q1949X/Q1949X^). Similarly, the tumor-prone resistance of DNA-damage was observed in the survival assays of *POLQ*^+/R1949X^ 293T and HCT116 cells (**Fig. 4**). Both 293T and HCT116 cell lines with p.Q1949X survived better under DNA damage stress induced by IR (**Fig. 4a**) or ETOPOSIDE (**Fig. 4b**). *POLQ*^+/Q1949X^ HCT116 cells have an increased POLQ protein level and a decreased γ-H2AX level under UV treatment (**Fig. 4c-e**), as mediated by enhanced TMEJ activity (**Fig. 4g-h**). These results demonstrated that PTC mutations lead to compensated overexpression of wild-type allele of POLQ under DNA-damage stress, which in turn causes overactivation of TMEJ pathway, resulting in enhanced error-prone DNA repair.

**Figure 4.**
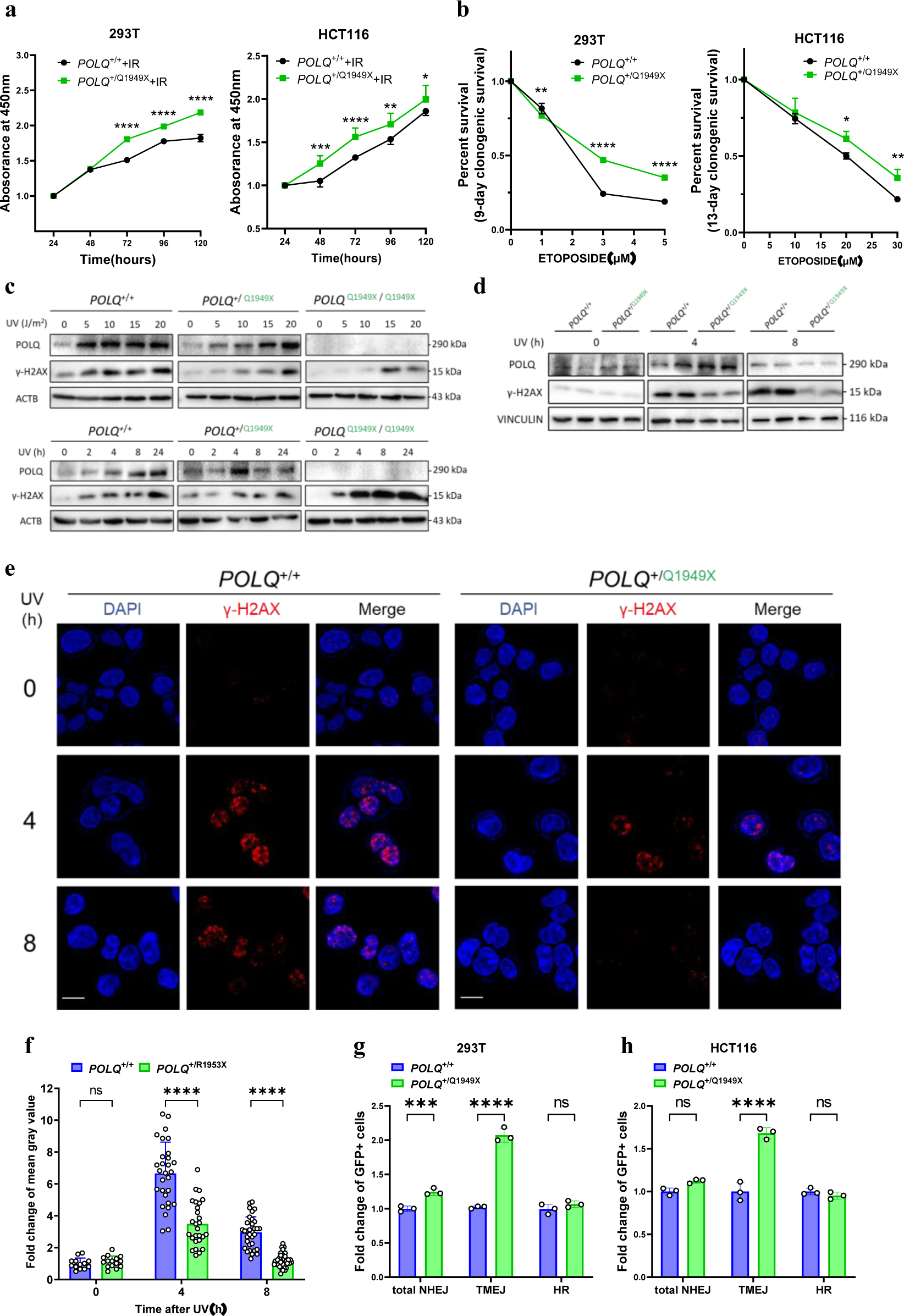
Introduction of another premature termination codon *POLQ* Q1949X mimic the R1953X-related cellular and molecular phenotypes. **a-b**, Both 293T and HCT116 cell lines with another heterozygous premature termination codon mutation (*POLQ*^+/Q1949X^ and *POLQ*^+/+^) survived better under DNA damage stress induced by 6 Gy IR (**a**) or ETOPOSIDE (**b**, 0-5uM for 293T and 0-30uM for HCT116). **c-d,** *POLQ*^+/Q1949X^ HCT116 cells have increased POLQ protein level and decreased γ-H2AX level under UV treatment. γ-H2AX and POLQ levels were measured by western blot analysis in cells 4h and 8h after UV release. VINCILIN, TUBULIN and ACTB were used as the loading control. **e**, Panels show images of γ-H2AX foci in cells at various time points under UV treatment, with γ-H2AX foci quantified in (**f**). Scale bars, 20µm. **g-h**, Percentage of GFP-positive cells are shown as pathway efficiency. Flow cytometric analysis of total-NHEJ, TMEJ and HR efficiency in *POLQ*^+/Q1949X^ and *POLQ*^+/+^ 293T (**g**) and HCT116 cells (**h**). *P*-value was determined by two-way analysis of variance. Data are shown as mean ± SD from at least three independent biological replicates. *P*-value was determined by two-way ANOVA. \**P* < 0.05; \*\**P* < 0.01; \*\*\**P* <0.001; \*\*\*\**P* < 0.0001 and ns, not significant.

### Targeting the TMEJ pathway reverses the POLQ PTC-mutation associated tumorigenesis-relevant phenotypes

Considering the fact that POLQ-PTC mutation carriers might be more resistant to DNA damage, it’s thus challenging for the treatment of those patients with the mutation. ETOP treatment inhibited the growth of *POLQ*^+/+^ cells well whereas *POLQ*^+/Q1949X^ cells showed a resistance to the drug treatment (**Fig. 1k-m**). We speculated that inhibition of the TMEJ by the FDA-approved antibiotic Novobiocin (NVB), a POLQ inhibitor, might have beneficial effects. As assumed, the reporter assays revealed that TMEJ activity could be suppressed by NVB, irrespective of the *POLQ* genotypes (**Fig. 5a**). Intriguingly, total NHEJ and HR in *POLQ*^+/+^ cells displayed increased activity after NVB treatment, which were not evident in *POLQ*^+/R1953X^ cells (**Fig. 5a**). These findings lend credence to the hypothesis that NVB may reverse the tumor phenotype of the POLQ mutant cells. Moreover, corroborating evidence emerged from the substantial upsurge in γ-H2AX expression levels induced by UV (10J/m2) following NVB treatment in *POLQ*^+/R1953X^ 293T cells (**Fig. 5b**). This outcome was further verified by immunofluorescence detection of γ-H2AX foci (**Fig. 5c**).

**Figure 5.**
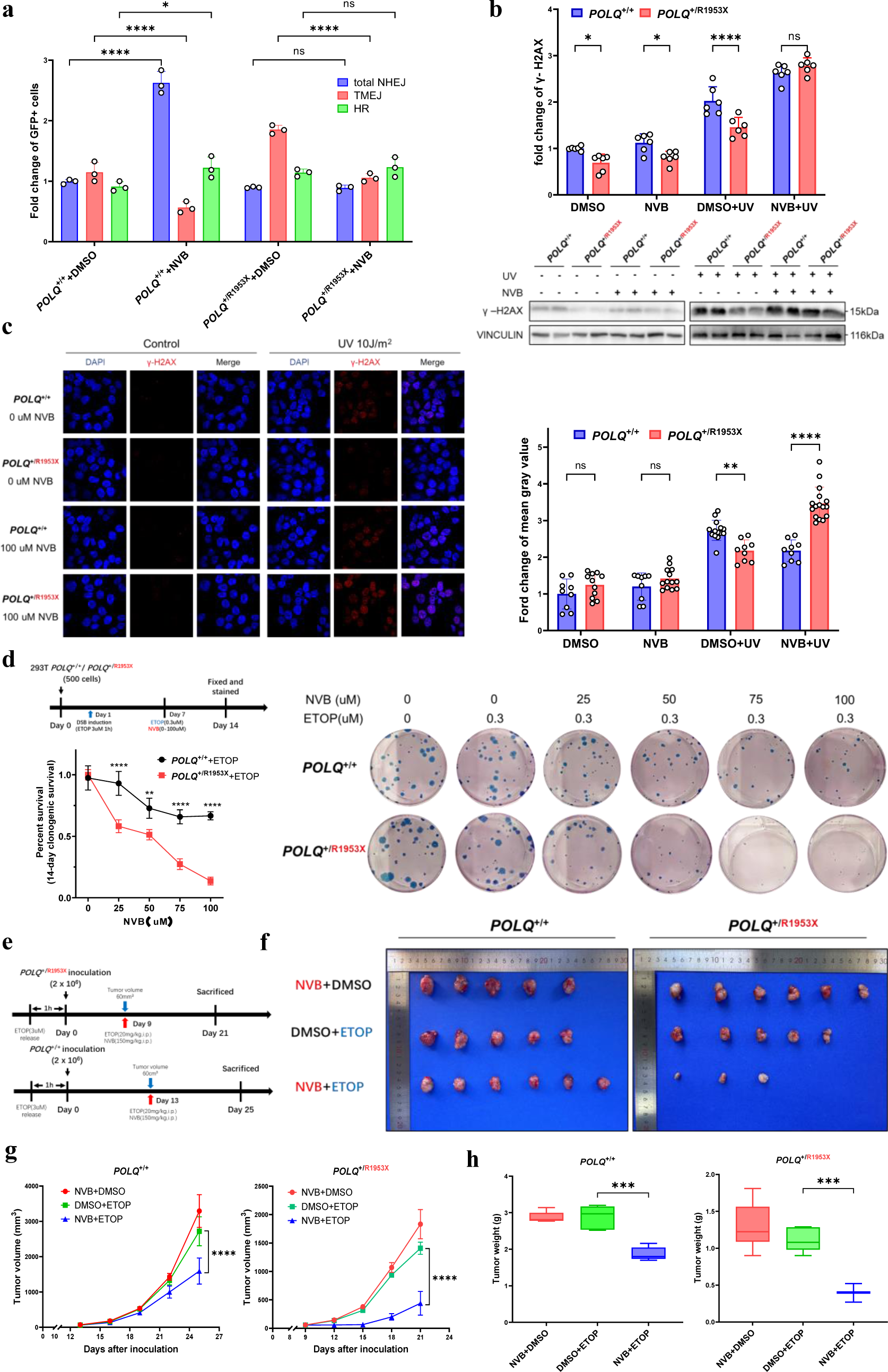
Modulation of TMEJ pathway reverses the POLQ-R1953X-related phenotypes. **a**, Novobiocin (NVB, 100uM), an inhibitor of POLQ, effectively inhibits the activated TMEJ efficiency of *POLQ*^+/R1953X^ compared with *POLQ*^+/+^ cells. The percentage of GFP-positive cells was used to reflect the efficiency of the repair pathway. *P*-value was determined by one-way analysis of variance. **b-c**, NVB can reverse the resistance of *POLQ*^+/R1953X^ cells to DNA damage. (b) *POLQ*^+/R1953X^ and *POLQ*^+/+^ cells were treated with DMSO/NVB 24 hours before UV-induced damage, cells were collected 2 hours after induction, and the expression level of DNA damage marker H2AX was detected by WB. (c) Immunofluorescence shows the display of UV-induced H2AX foci under DMSO/NVB treatment. Scale bar, 20µm. Statistics are on the right. *P*-value was determined by one-way analysis of variance. **d**, Colony formation experiments suggest that NVB can reverse the abnormal proliferation ability of *POLQ*^+/R1953X^ cells under DNA damage stress. *P*-value was determined by one-way analysis of variance. **e-h**, NVB inhibits the in vivo growth of *POLQ*^+/R1953X^ cell xenografts after DNA damage induction. (**e**) Diagram shows the experimental plan of xenografting of DNA damage-induced 293T cells in BALB/c nude mice, and the treatment with ETOPOSIDE and NVB. *POLQ*^+/R1953X^ and *POLQ*^+/+^ 293T cells were treated *in vitro* with ETOPOSIDE or DMSO for 1 hour, then xenografted into BALB/c nude mice. When the xenograft grew to 0.06cm^3^, intraperitoneal injection of ETOPOSIDE with or without NVB was performed. Tumor samples were collected 12 days after injection. (f) Shows the display of the xenograft. Statistical analysis of tumor volumes (g) and tumor weights (h) in different groups(n=6/group). *P*-value was determined by Student’s *t*-test. Data are shown as mean ± SD. \**P* < 0.05; \*\**P* < 0.01; \*\*\**P* <0.001; \*\*\*\**P* < 0.0001 and ns, not significant.

We next explored whether NVB could counteract DNA damage resistance in *POLQ* heterozygous mutant cells. *POLQ*^+/R1953X^ and *POLQ*^+/+^ 293T cells were exposed to NVB under DNA-damage stress. NVB was found to significantly reverse resistance to UV-induced DNA damage in *POLQ*^+/R1953X^ cells compared to *POLQ*^+/+^ cells (**Fig. 5d**). We subsequently conducted 293T xenograft experiments, which showed that ETOPOSIDE induced substantial amount of DNA damage to *POLQ*^+/R1953X^ xenografts when mice were administered with NVB (**Fig. 5e-h and Supplementary Fig. S3a-d**). In line with prior findings, xenografts expanded more rapidly and extensively in mice inoculated with DNA-damage-induced *POLQ*^+/R1953X^ 293T cells. The DNA-damage potency of ETOPOSIDE was significantly amplified in NVB-treated *POLQ*^+/R1953X^ xenografts (**Fig. 5e-h and Supplementary Fig. S3a-d**).

We also examined the potential of NVB for increasing the radiosensitivity of *POLQ*^+/R1953X^ HCT116 cells. The results suggested that NVB could significantly inhibit the radiotherapy resistance of *POLQ*^+/R1953X^ HCT116 xenografts and improved the radiotherapy sensitivity (**Supplementary Fig. S3e-h**). These observations provided compelling evidence that NVB counteracts DNA damage resistance resultant from the POLQ mutation by impeding the TMEJ pathway, further strengthened the evidence that overactivation of TMEJ is the underlying mechanism enhancing DNA repair in cells with POLQ heterozygous PTC mutations.

## Discussion

For the first time, we demonstrated that the germline POLQ p.Arg1953X mutation predisposes individuals to hereditary colorectal adenomatous polyposis and CRC. This allele exhibits dominant inheritance, with apparent high penetrance, and the related tumors predominantly arise in the colorectum. Some individuals display a multiple-adenoma phenotype, resembling the *MUTYH*-associated polyposis (MAP), a condition arising from defective base excision repair. Others with the p.Arg1953X mutation developed early-onset carcinoma in the colorectum, endometrium, or other extracolonic organs, mirroring Lynch syndrome, which results from defective DNA mismatch repair. The disparate tumor spectrum among carriers of the POLQ p.Arg1953X remains unexplained. Similar differences are observed in polymerase proofreading-associated polyposis (PPAP), where carriers of the POLD1 mutations seem to be relatively prone to endometrial cancer^7^. We observed that five affected women developed cancerous or precancerous lesions (complex hyperplasia) in the endometrium. Consequently, colorectal tumor presence cannot be considered as an obligatory requisite to define this syndrome. Analyzing only the colorectal phenotype, individuals with the Arg1953X mutation exhibited two phenotypes: multiple adenomas and early-onset colorectal cancer. This phenotypic variation may be impacted by additional genetic modifiers^17^ or different environments and lifestyles.

How the POLQ PTC mutation gives rise to the tumor-related phenotypes is a pivotal question both for understanding the mechanism and for treatment. Three hypotheses have been proposed: (1) the presence of a deleterious truncated peptide, (2) *POLQ* haploinsufficiency, and (3) a novel hybrid mechanism. Using genome editing technologies, our study explored the mechanisms underlying the tumorigenesis associated with POLQ PTC mutations *in vitro* and *in vivo*. Our results demonstrated that cells possessing a heterozygous POLQ mutation exhibit abnormal proliferation and apoptosis patterns under replication or DNA damage stress. This occurs without any evidence of *POLQ* haploinsufficiency at the protein level and/or in the absence of detectable truncated peptides in cell lines carrying two separate POLQ PTC mutations. These findings suggested a potentially novel pathogenic mechanism other than haploinsufficiency or toxic truncated peptides. In fact, we identified a unique molecular signature characterized by the activation of the nonsense-mediated mRNA decay (NMD) pathway and dysregulation of POLQ wild-type allele expression. The NMD pathway has been identified as the primary mechanism responsible for the clearance of mRNA with PTC mutations^18^. However, in addition to its function as a cellular surveillance pathway, the degradation of mRNA also initiates genetic compensation, a process integral to maintaining functional homeostasis^19^. Our study revealed that DNA damage triggers a compensatory upregulation of the *POLQ* wild-type allele. Intriguingly, we discovered that this compensatory expression of *POLQ* wild-type allele not only maintains DNA repair functionality but also exhibits a strikingly abnormal enhancement.

Cancer typically develops through multiple steps, each involving the clonal evolution of cells with abnormal defense mechanisms^20^. In our study, we found that microsatellite instability (MSI)/hypermutation could underlie this stepwise progression. How does genetic compensation of POLQ PTC mutation lead to the MSI/hypermutation phenotype? Our data indicated that compensated expression of *POLQ* in mutant cells triggers an overactivation of the error-prone POLQ-mediated end joining (TMEJ) pathway. Consequently, repair via the error-prone TMEJ pathway contributes to the accumulation of detrimental mutations, leading to the hypermutation phenotype^21^. When treated with *POLQ* inhibitor NVB ^16^, *POLQ* mutation-induced TMEJ pathway overactivation could be mitigated. This subsequently heightened the sensitivity of the mutant cells to DNA-damage agents, a phenomenon not observed in wild-type cells. This hints at the critical role of the TMEJ pathway in preserving DNA repair functionality in POLQ mutant cells.

Interestingly, studies have demonstrated that in MMR-deficient cells, replication stress-associated double-strand breaks (DSBs) are repaired by TMEJ in tandem with the induction of MSI, rather than by the MMR-proficient cells-dependent NHEJ pathway, resulting in chromosomal instability ^12,22^. It remains a mystery, however, as to how MSI is specifically triggered in a POLQ-mutant and MMR-proficient context, but it is likely associated with TMEJ overactivation. In conclusion, for cells harboring a POLQ PTC mutation, a high activation level of the TMEJ pathway could be the key for the MSI/hypermutation phenotype, with NVB showing the potential as a therapeutic strategy for the treatment of POLQ-mutant tumors. These speculations were supported by a recent study demonstrating that NVB eliminates HR-deficient tumor cells via selective TMEJ pathway inhibition both *in vitro* and *in vivo*^16^.

The POLQ mutation underscores the pivotal role of double-stranded DNA break repair in predisposing individuals to colorectal, endometrial, breast, and ovarian cancers. Individuals with a family history of multiple colorectal adenomas and/or multiple or early-onset colorectal (or endometrial) carcinoma that can’t be fully explained by known pathogenic genes should be detected for the *POLQ* gene, especially PTC mutations like p.Arg1953X.

In short, our study defines a new *POLQ*-type CRC, provides insights into how POLQ mutations lead to distinct tumor phenotypes, and suggests for potential therapeutic strategies. Our findings support the crucial role of POLQ in safeguarding genome integrity and its involvement in the predisposition to colorectal and endometrial cancers. Future efforts are warranted to broaden our understanding of *POLQ*-mediated tumorigenesis and therapeutic target for these cancers.

## Online Methods

### Subjects

The subjects in this study are all Han Chinese resident in Yunnan Province, China. All studies were conducted with the approval of the ethics committee of Kunming Medical University (2017-3), and all participants provided written informed consent. During the discovery phase, families were recruited from a single medical institution. We selected probands predicted to have high phenotypic penetrance. Specifically, we focused on those with multiple tumors, an early age of onset, and multiple family members diagnosed with colorectal cancer (CRC) or adenomas. Owing to the complex nature of these clinical characteristics, we implemented broad inclusion criteria. After a thorough screening, we identified a set of 12 families. In all these families, probands were diagnosed with CRC, and there was at least one first-or second-degree relative with CRC. All probands had been evaluated for Lynch syndrome through examination of mismatch repair protein expression in tumors and/or by direct testing for *MSH2, MLH1, MSH6*, and *PMS2* mutations. We subsequently performed whole-exome sequencing on peripheral blood DNA from these probands, as well as at least two affected and one healthy relative.

In the verification phase, we recruited 84 families from 5 medical institutions, following the same selection criteria. We also collected 310 cases of sporadic CRC in the verification phase. Alongside these newly recruited family probands, the POLQ Arg1953X mutation was confirmed through Sanger sequencing. All the cases were Han Chinese residents of Yunnan Province.

### Genotyping

The whole-exome sequencing data of *POLQ* were verified in LS7 family by Sanger sequencing using standard methods, while other members within the LS7 family were also genotyped. For the samples in the verification phase, we used Sanger sequencing to detect the POLQ Arg1953X mutation and included one known mutation and one wild-type sample as the control in each run. For the LS13 family with the POLQ Arg1953X mutation found in the verification phase, Sanger sequencing of other members of the family was performed. WES was used to exclude the known pathogenic gene mutations in this family.

### Analysis of the TMB and MSI status from NGS data

The TMB (mutations per megabase (Mb) of DNA) was extrapolated using sequencing data from the panel of 769 cancer-related genes and determined by analyzing the number of somatic mutations per megabase. The top 25^th^ percentile of the TMB value was used as the cutoff value to define tumors with a high mutation burden (TMB-H tumors) in this study.

Tumor DNA samples were subjected to NGS using the cancer gene-targeted panel. Fifty-five target microsatellite loci were examined and compared with those in genomic data from healthy people in the Chinese database. The number of microsatellite loci altered by somatic insertions or deletions was determined for each patient sample. If the ratio of unstable loci to passing loci was equal to or higher than 0.15, the MSI status of the sample was defined as MSI-H; if the ratio of unstable loci to passing loci was less than 0.15, the MSI status of the sample was defined as MSI-L/MSS.

### Cell line and reagents

The primary antibodies and chemicals used in this study are listed in **Supplementary Table 5**. The HEK293T cells were obtained from the Kunming Cell Bank, Kunming Institute of Zoology (KIZ), and were cultured in Dulbecco’s modified Eagle’s medium (DMEM; Thermo Fisher) supplemented with 10% fetal bovine serum (FBS; Thermo Fisher) at 37 °C in 5% CO_2_. ETOPOSIDE and Novobiocin were dissolved in dimethyl-sulfoxide (DMSO) at an appropriate concentration as a stock solution and stored at −80 °C before further use.

### Vector construction and transfection

Myc and Flag-tagged POLQ expression plasmids were purchased from Addgene (#73132). Site-directed mutagenesis of Arg1953X was performed using a pair of primers (**Supplementary Table 6**) designed to create a C>T mutation by using the Easy Mutagenesis System (Beijing TransGen Biotech) according to the manufacturer’s protocols. All constructs were confirmed by Sanger sequencing.

The transient transfection of vectors was performed using Lipofectamine™ 3000 (Invitrogen, L3000015) according to the manufacturer’s protocols. Briefly, cells were seeded on 6-well plate. When cells are 50%–60% confluent, growth medium was removed and cells were washed once with Opti-MEM medium (Gibco-BRL, 31985– 070) for transfection. Expression vectors or empty vector (2.5 μg/well) were dissolved in Opti-MEM medium (125 µL/well) containing P3000™ regent (5 μL/well), then were mixed with Lipofectamine™ 3000 (3.75 μL/well) diluted in Opti-MEM medium (125 μL/well). The mixture was incubated at room temperature for 15 min and added to each well together with an additional Opti-MEM medium (750 µL/well). The medium was removed at 6h after transfection and fresh growth medium (2 mL/well) was added for growth until harvest at 48 h after transfection.

### Genome editing of HEK293T and HCT116 cell lines

To generate isogenic cell lines encoding Arg1953X in *POLQ*, we employed a CRISPR/Cas9-based point mutation knock-in approach following the procedure in our previous studies. This was executed by transfecting vector px330-mcherry (Addgene plasmid #98750), post-ligation of two annealed reverse complementary guide DNA oligos to initiate site-specific DNA double-strand breaks (**Supplementary Fig. 2a**). We determined the guide RNA according to the mutation position (**Supplementary Table 6**). We designed donor single-stranded oligo-DNAs (ssODN) for homologous recombination to mutate the specific locus of *POLQ* (**Supplementary Table 6**). We co-transfected cell lines with the CRISPR/Cas9 vector and ssODN using Lipofectamine 3000 (Thermo Fisher Scientific), applying 2.5 µg of the CRISPR/Cas9 vector and 5 µg of ssODN to each well of a 6-well plate containing 60% confluent cells. After 48 hours post-transfection, we sorted the cells by flow cytometry for mcherry expression, collected positive cells, and seeded them at one cell per well in a 96-well plate for single-cell clone expansion. Two to three weeks after seeding, we prepared DNA from amplified single-cell clones for genotyping via Sanger sequencing.

We utilized base-editing to generate precise point mutations in cellular DNA for the control variants (Gln1949*) (**Supplementary** Fig. 2a). We use the same guide RNA (gRNA) targeting the genomic regions of Arg1953X and sub-cloned them into the pGL3-U6-sgRNA-PGK-puromycin plasmid (Addgene plasmid #51133). Constructs containing varying gRNAs (500 ng per well in a 6-well plate) were co-transfected with pCMV-BE4 (2 μg per well in a 6-well plate, Addgene plasmid #100802) into HEK293T cells using Lipofectamine 3000 (Thermo Fisher Scientific). Post-transfection (24h), we replaced the culture medium daily with fresh medium supplemented with 2 μg/mL puromycin, selecting cells with puromycin for five days. Single cells resistant to puromycin were sorted into a 96-well plate by flow cytometry and cultured for 2-3 weeks to obtain single-cell clones. For each clone, we amplified and sequenced the target region to confirm successful editing of the target variants.

### Cell viability and colony formation assays

Cell viability in 96-well plates with different treatment was measured using the Cell Counting kit-8 (Beyotime C0038). Briefly, cells were seeded in 96-well plates at 1000cells/well in 100μL of standard medium. Then, 10 μL of CCK8 substrate was added to each well on the 0, 1, 2, 3, 4 and 5 days after DNA damage agent induction, and incubated in the dark for 2h. The absorbance value (OD) at 450 nm was measured with a full wavelength scanner.

Cells were seeded in 6-well plates at 500cells/well in 2mL of standard medium. Colony formation ability in complete growth medium after different treatment was determined by using ImageJ software to count the number of cells, which were fixed with methanol and stained with modified giemsa staining solution (Beyotime C0131). Survival fraction of cells was the ratio of the plating efficiency of treated cells to that of control cells.

### DNA synthesis assay

To detect the DNA synthesis of CRC cells, we used the BeyoClick™ EdU Cell Proliferation Kit with Alexa Fluor 647 (Beyotime C0080L) according to the manufacturer’s protocol. Briefly, cells were seeded in 12-well plates at 1×10^4^ cells/well in 1mL of standard medium, alone or treated with IR (6Gry). After 24 h, the cells were incubated with 10 μM EdU in conditioned medium for 2 h, followed by fixing, permeabilizing, and staining. For each sample, at least ten random fields were observed using fluorescence microscopy, and mean gray values were counted.

### Apoptosis assay by flow cytometry

An Annexin V FITC/PI (556547, BD, USA) apoptosis staining assay was performed to detect cell apoptosis. In brief, 2×10^5^ cells were collected using trypsin without EDTA. After two washed with PBS, cells were incubated in the dark with 500 μL of Binding buffer, 5 μL of AnnexinV-FITC and 10 μL of PI for 10 min at room temperature. The fluorescence intensity of samples was quantified by flow cytometry (BD FACSCanto II; BD Biosciences, CA, USA). Experiments were carried out three times.

### Cell-cycle analysis

The cells were seeded in 12-well plates at 50%–60% confluence and treated as indicated 24 h after plating. Then, the cells were harvested at 0, 6 and 24 h after DNA damage induction. After having been fixed in 70% ice-cold ethanol overnight, cells were then stained with propidium iodide in the presence of RNase A (Beyotime C1052). Fluorescence intensity was measured by flow cytometry. The percentages of cells in G0/G1, S, and G2/M phases were analyzed in Flowjo software.

### Xenograft experiments

Five-week-old female BALB/c nude mice were purchased from Vital River Laboratory (Beijing, China) and housed in flow cabinets under specific pathogen-free conditions. Animal feeding and experiments were approved by the animal ethics committee of Kunming Institute of Zoology, Chinese Academy of Sciences. After a short period of adaptation, mice were randomly subjected to four groups (n =6 per group). Food and water were supplied *ad libitum*. Then, 293T (2×10^6^) or HCT116 (1×10^6^) cells were injected subcutaneously into the axilla of each mouse to establish the xenograft model. Tumor volumes and mouse weights were measured every 3 days. For Etoposide, IR, and Novobiocin treatment, mice were randomly re-divided into three groups for each genotype when the tumor volume was close to 60 mm^3^, 120 mm^3^ or 1500 mm^3^. With IR release (10Gy), we conducted intraperitoneal injection of Etoposide (20 mg/kg) and Novobiocin (150 mg/kg) alone or in combination. Etoposide and Novobiocin were dissolved in dimethyl sulfoxide and then diluted by normal saline.

### Quantitative real-time PCR

Total RNA was isolated from cell using TRIZOL (Invitrogen, 15596-018). Total RNA (1 μg) was used to synthesize single-strand cDNA using the M-MLV Reverse Transcriptase (Promega, M170A) in a final volume of 25 μL according to the manufacturer’s instructions. The relative mRNA levels of *POLQ* were quantified by using quantitative real-time PCR, with normalization to the Actb/β-actin gene. The quantitative real-time PCR was performed in a total volume of 20 μL containing 2 μL of diluted products, 10 μL of SYBR Master Mix (BioRad), 0.2 uL 10 μM each primer (**Supplementary Table 5**), on a BIO-RAD Real-time PCR detection system. The quantitative real-time PCR thermal cycling conditions were composed of a denaturation cycle at 95 °C for 5 min, followed by 40 cycles of 95 °C for 10 s and 55 °C for 30 s.

### Immunofluorescent staining of γ-H2AX

Seeded on coverslips to 60-70% confluency, cells were allowed to attach for 12 h and then treated with Etoposide or ultraviolet. At the time points as indicated in corresponding figures, cells were washed with PBS and fixed in 4% paraformaldehyde for 8 min. After three washings, cells were permeabilized with 0.2% Triton X-100. The coverslips were then blocked with 1% BSA in PBS containing 0.1% Triton X-100 and stood for 30 min at room temperature. The blocked coverslips were then probed with antibody against γ-H2AX, followed by Alexa Fluor Plus 594 secondary antibody and DAPI (**Supplementary Table 5**). Images of fluorescent γ-H2AX foci was captured by confocal microscopy.

### Western blotting

Cells were lysed with SDS lysis buffer (Beyotime, P0013) containing PMSF (Beyotime, ST506), phosphatase inhibitor cocktail A (Beyotime, P1081). The protein concentration of each sample was determined by BCA assay using the BCA kit (Beyotime, P0011). Lysate containing 50-80µg of protein was separated on 8% or 12% SDS PAGE and transferred to PVDF membranes (BioRad L1620177 Rev D). The membrane was soaked with 5% (w/v) skim milk for 2 h at room temperature, then was incubated with primary antibody (**Supplementary Table 5**) overnight at 4 ℃. The membrane was washed 3 times with TBST (Tris-buffered saline [Servicebio, G0001] with 0.1% Tween 20 [Sigma, P1379]), each time 5 min, followed by incubation with the secondary antibody. Peroxidase-conjugated anti-mouse (lot number 474–1806) or anti-rabbit (lot number 474–1516) IgG (1:10000; KPL) were used as the respective secondary antibodies. The epitope was visualized using an ECL Western Blot Detection Kit (Millipore, WBKLS0500). We used ImageJ (National Institutes of Health, Bethesda, Maryland, USA) to evaluate densitometry of each blot.

### GFP reporter-based DNA repair assays

The GFP reporter-based DNA repair assays were performed as previously described ^1^. In brief, DR-GFP (for HR), EJ5 (for total NHEJ) and EJ2 (for TMEJ) repair substrates were purchased from Addgene (#26475, #44026, and #44025). To measure repair efficiency, 1×10^5^ cells were plated in each well of a 12-well plate. Indicated compounds or DMSO were added to the medium for 24 h, then cells were transfected with the repair substrate and I-SceI enzyme in equal proportion. 48 hours after transfection, cells were trypsinized, and GFP-positive cells were quantified by flow cytometry (Beckman).

## Data availability

All data generated or analyzed during this study are included in this article and its supplementary files.

## Acknowledgments

The study was supported by the National Natural Science Foundation of China (31660312 and 82160563) and the Applied Basic Research Foundation of Yunnan Province. The funders had no role in study design, data collection and analysis, decision to publish, or preparation of the manuscript.

