## Supplementary figures for "A germline heterozygous *POLQ* nonsense mutation causes hereditary colorectal cancer"

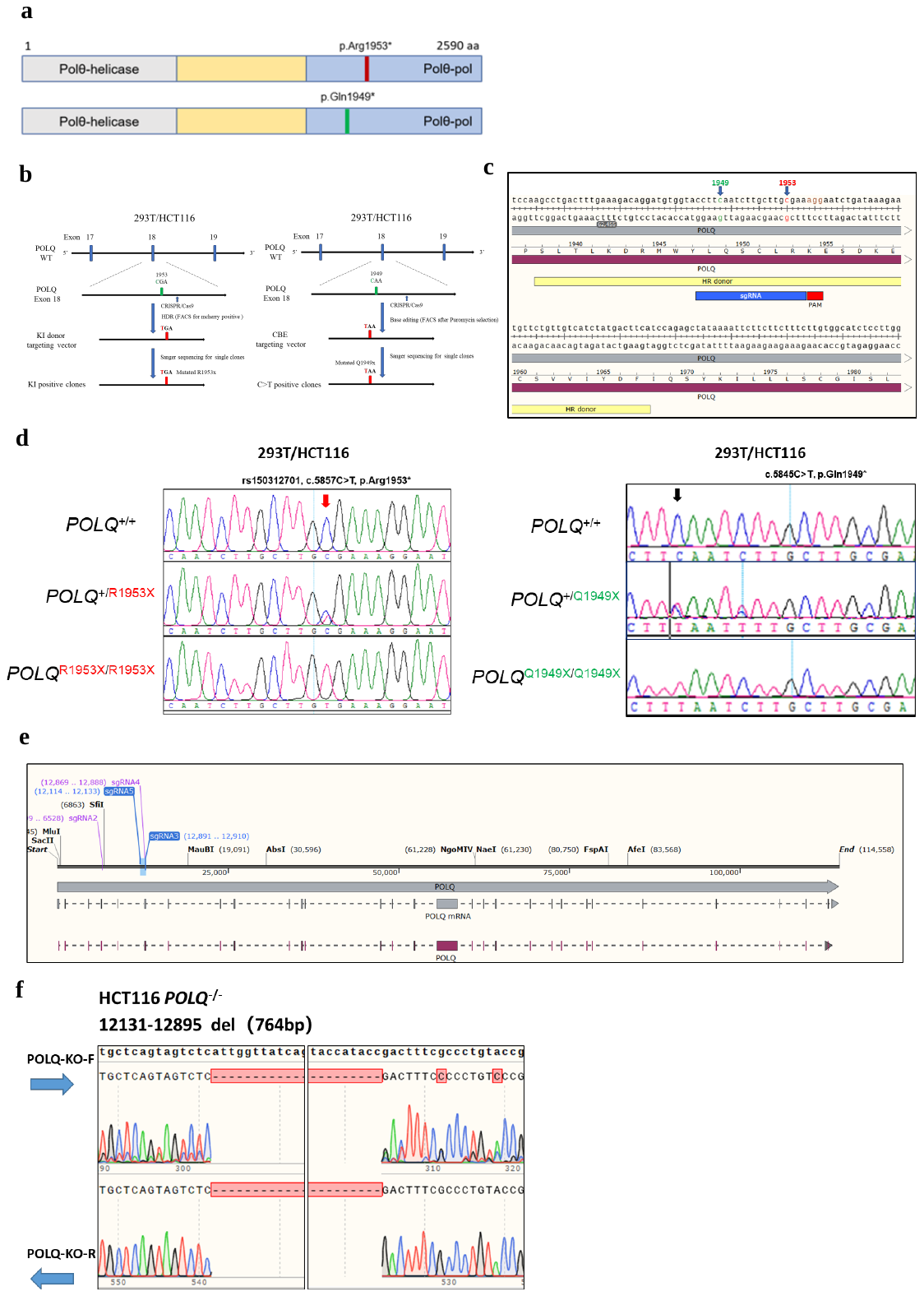

**Supplementary Figure S1**. **Genome editing of 293T and HCT116 cell lines. a**, Locations of PTC mutations in *POLQ* DNA and protein resulting in premature stop codons (PTC). **b,** CRISPR/Cas9-based strategy in the example to introduce the Arg1953* and Gln1949* mutation in wildtype HEK293T and HCT116 cell lines knock-in and base-editing vector systems. **c,** CRISPR/Cas9 based strategy. Guide-RNA targeted sequence is marked in blue, the protospacer adjacent motif (PAM) is marked in red, and the sequence of the single-stranded DNA oligo is marked in yellow. **d,** Sanger sequencing traces of heterozygous and homozygous mutated cell lines. Point mutation is marked with arrow. **e-g**, Establishing *POLQ* Knock-out（*POLQ*^-/-^）HCT116 cell line based on CRISPR/Cas9 system. (**e**) The double sgRNA(Supplementary Table S6) targeting sequences of the *POLQ* gene were designed by using the CRISPOR website (http://crispor.tefor.net/). Two sgRNA were designed to target c.248-285 in the *POLQ* gene. (**f**) Sanger sequencing chromatograms showed the sgRNA-targeted region in HCT116 knockout cell clones. The NM_199420.4 of the *POLQ* gene was used as the reference sequence (Ref-Seq).

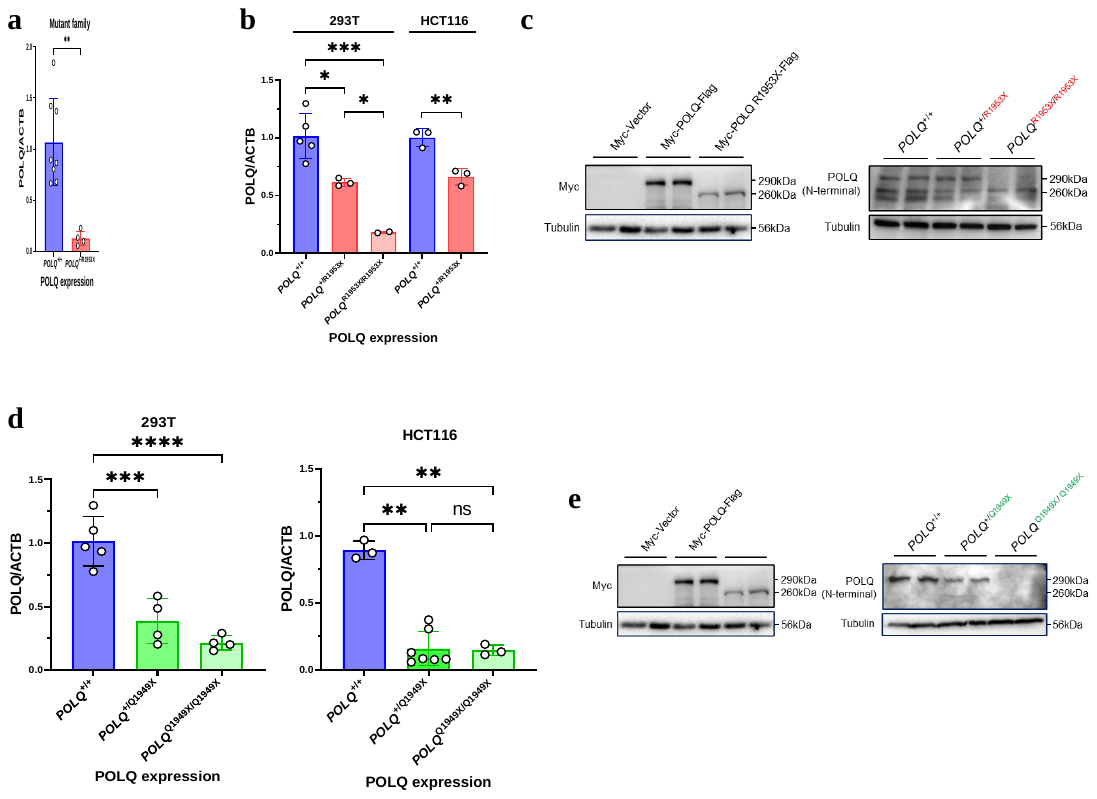

**Supplementary Figure S2**. **mRNA and protein level of POLQ with mutations.**

**a**, Detection of POLQ mRNA expression in blood of 7 healthy individuals and 4 *POLQ* mutant patients from two positive families measured by qPCR and verified in isogenic 293T and HCT116 cell lines (**b, d**). *P*-value was determined by Student’s t-test.

**c** and **e**, No truncated *POLQ* peptide was detected in homozygous mutant 293T cells (*POLQ*^R1953X/R1953X^).

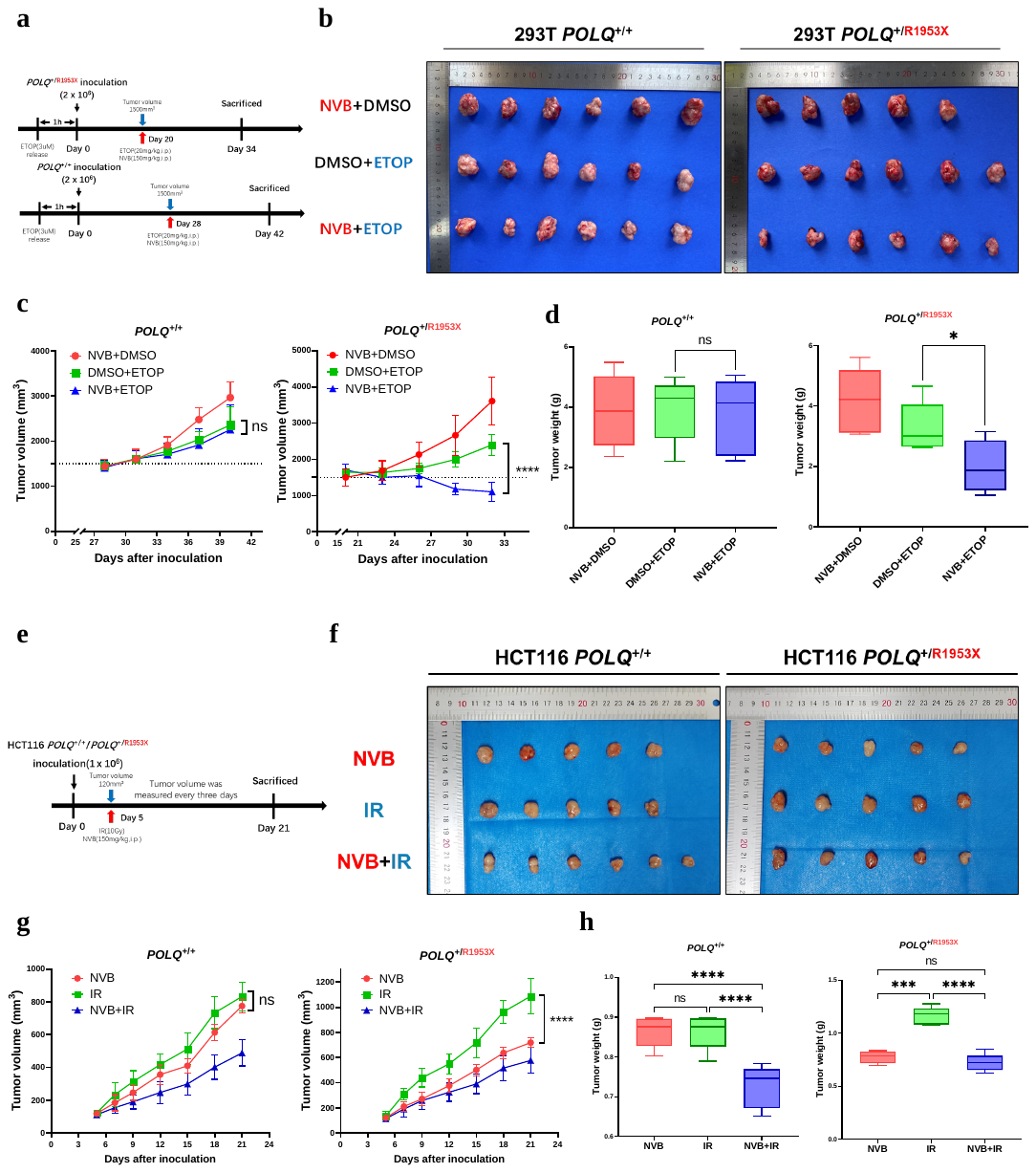

**Supplementary Figure S3**. **NVB inhibits the in vivo growth of *POLQ*^+/R1953X^ cell xenografts. a-d** NVB can inhibit the growth of *POLQ*^+/R1953X^ 293T cell xenografts induced by DNA damage. (**a**) Diagram shows the experimental plan of xenografting of DNA damage-induced 293T cells in BALB/c nude mice, and the treatment with ETOPOSIDE and NVB. *POLQ*^+/R1953X^ and *POLQ*^+/+^ 293T cells were treated in vitro with ETOPOSIDE or DMSO for 1 hour, then xenografted into BALB/c nude mice. When the xenograft grew to 0.06cm^3^, intraperitoneal injection of ETOPOSIDE with or without NVB was performed. Tumor samples were collected 14 days after injection. **(b)** Shows the display of the xenograft. Statistical analysis of tumor volumes (**c**) and tumor weights (**d**) in different groups(n=6/group). *P*-value was determined by Student’s *t*-test. **e-h** Injection of NVB showed an increase sensitivity of *POLQ*^+/R1953X^ HCT116 cell xenografts to IR. (**e**) Diagram shows the experimental plan of xenografting of HCT116 cells in BALB/c nude mice, and the treatment with IR and NVB. *POLQ*^+/R1953X^ and *POLQ*^+/+^ HCT116 cells were xenografted into BALB/c nude mice. When the xenograft grew to 0.12cm^3^, IR(10Gy) and intraperitoneal injection NVB(150mg/kg) were performed. Tumor samples were collected 16 days after injection. (**f**) Shows the display of the xenograft. Statistical analysis of tumor volumes (**g**) and tumor weights (**h**) in different groups(n=6/group). *P*-value was determined by Student’s *t*-test. Data are shown as mean ± SD. **P* < 0.05; ***P* < 0.01; ****P* <0.001; *****P* < 0.0001 and ns, not significant.
